# Molecular stratification of endometrioid ovarian carcinoma predicts clinical outcome

**DOI:** 10.1101/2020.03.19.20038786

**Authors:** Robert L Hollis, John P Thomson, Barbara Stanley, Michael Churchman, Alison M Meynert, Tzyvia Rye, Clare Bartos, Yasushi Iida, Ian Croy, Melanie Mackean, Fiona Nussey, Aikou Okamoto, Colin A Semple, Charlie Gourley, C. Simon Herrington

**Author notes:** **Corresponding Author:** Professor C Simon Herrington - Nicola Murray Centre for Ovarian Cancer Research, Cancer Research UK Edinburgh Centre, MRC IGMM, University of Edinburgh. Crewe Road South, Edinburgh, EH4 2XU, UK. These authors contributed equally (Robert L Hollis, John P Thomson, Barbara Stanley, Charlie Gourley, C. Simon Herrington). **Conflicts of interest:** MM: honoraria from Tesaro, BristolMyersSquibb and Roche. FN: non-personal interests in AstraZeneca and Tesaro. CG discloses: research funding from AstraZeneca, Aprea, Nucana, Tesaro and Novartis; honoraria/consultancy fees from Roche, AstraZeneca, MSD, Tesaro, Nucana, Clovis, Foundation One, Cor2Ed and Sierra Oncology; named on issued/pending patents related to predicting treatment response in ovarian cancer outside the scope of the work described here. All other authors declare no conflicts of interest.

## Abstract

Endometrioid ovarian carcinoma (EnOC) is an under-investigated type of ovarian carcinoma. Here, we report the largest genomic study of EnOCs to date, performing whole exome sequencing of 112 cases following rigorous pathological assessment. High frequencies of mutation were detected in *CTNNB1*(43%), *PIK3CA*(43%), *ARID1A*(36%), *PTEN*(29%), *TP53*(26%) and *SOX8*(19%), a novel target of recurrent mutation in EnOC. *POLE* and mismatch repair protein-encoding genes were mutated at lower frequency (6%, 18%) with significant co-occurrence. A molecular taxonomy was constructed using a novel algorithm (PRISTINE), identifying clinically distinct EnOC subtypes: *TP53*m cases demonstrated greater genomic complexity, were frequently FIGO stage III/IV at diagnosis (48%) and incompletely debulked (44%), and demonstrated inferior survival; conversely, *CTNNB1*m cases demonstrated low complexity and excellent clinical outcome, were predominantly stage I/II at diagnosis (89%) and completely resected (87%). Tumour complexity provides further resolution within the *TP53*wt/*CTNNB1*wt group. Moreover, we identify *t*he WNT, MAPK/RAS and PI3K pathways as good candidate targets for molecular therapeutics in EnOC.

## Introduction

Ovarian carcinomas (OC) are a heterogeneous group of malignancies comprising 5 core histological types, each with distinct pathological characteristics, molecular landscapes and clinical behaviour^1, 2^. Endometrioid OC (EnOC) accounts for approximately 10% of all OCs, with the majority of cases diagnosed as low grade, early stage disease with excellent clinical outcome^3, 4^.

Currently, the management of EnOC follows the historic ‘one size fits all’ approach of aggressive cytoreductive surgery with adjuvant platinum-taxane chemotherapy for patients with disease that has progressed beyond the ovary/fallopian tube. By contrast, routine molecular stratification of care is emerging in other OC types, most notably with the advent of poly(adenosine diphosphate-ribose) polymerase (PARP) inhibitor therapy in *BRCA1/2*-mutant high grade serous (HGS) OC cases^5, 6^.

Targeted sequencing approaches have identified *PTEN, ARID1A, PIK3CA, KRAS, CTNNB1* and genes encoding mismatch repair (MMR) proteins as frequently mutated in relatively small cohorts of EnOC^7, 8, 9^, reminiscent of endometrioid endometrial carcinomas (EnECs)^10^, with a *TP53* mutation rate markedly lower than their high grade serous (HGS) OC counterparts^11^. Recent whole genome sequencing of a small EnOC case series has recapitulated these findings and identified a small proportion of EnOC with extensive copy number aberrations more akin to the genomic instability demonstrated by HGSOCs^12^.

The majority of EnOC are believed to arise from endometriosis^1^, and most grade 1 and 2 (low grade) EnOC display a classical immunohistochemical (IHC) profile comprising WT1 negativity, wild-type p53 expression, and oestrogen receptor (ER) positivity^4^. These classical low grade EnOC bear close histological resemblance to EnECs^13^.

Grade 3 (high grade) EnOC can be challenging to differentiate from HGSOC on the basis of morphology alone^4, 13^. In particular, HGSOCs demonstrating the <u>s</u>olid, pseudo-<u>e</u>ndometrioid and/or transitional-cell-like (SET) histological pattern, which may be associated with *BRCA1* mutations^14^, represent a population easily misclassified as EnOC. Indeed, it is now recognised that many historically diagnosed high grade EnOCs in fact represent HGSOCs, supported by transcriptomic studies demonstrating that a proportion of high grade EnOCs cluster with HGSOCs^15, 16, 17, 18^. As such, true high grade EnOC are increasingly rare, representing only around 5-19 % of EnOC cases^4, 8, 19, 20^; these cases reportedly experience poor clinical outcome, in contrast to their low grade counterparts^3, 20^.

To date, the overwhelming body of clinical and molecular EnOC characterisation has been confounded by the inclusion of historically misclassified HGSOCs. Mutational analyses performed by more recent studies have either been applied to low grade EnOC alone^21^, or lack information on grade or diagnostic criteria used^7, 22^ and have ubiquitously analysed small patient cohorts with insufficient power to confidently correlate molecular events with patient outcome. As a result, the molecular landscape of EnOC, in particular high grade EnOC, is poorly defined.

WT1 immunohistochemistry (IHC) is a useful tool to discriminate high grade EnOC (WT1 negative) from HGSOC (WT1 positive), reducing inter-observer variation^15, 19, 23, 24, 25, 26^. Here we perform molecular characterisation of contemporarily defined EnOC, with the use of IHC for WT1. We perform whole exome sequencing (WES) to define the genomic landscape of EnOC, including high grade EnOC, in a sizeable cohort of otherwise unselected patients and correlate specific molecular events with differential patient outcome.

## Results

### Clinical characteristics

Of 289 historically diagnosed EnOC cases identified with available tumour material, 112 WT1 negative cases were characterised by WES following rigorous pathology review (18 non-evaluable, 8 non-ovarian primary, 120 WT1 positive, 14 clear cell/mucinous/carcinosarcoma/mixed histology, 2 concurrent metastatic malignancy, 15 DNA extraction/WES failure) (figure 1). The clinicopathological characteristics of these patients are shown in table 1. The majority (78.2%, 86 of 110 evaluable cases; 2 unknown stage) of patients presented with stage I or II disease. 19 cases (17.0%) had synchronous endometrial cancer diagnosis. The median follow-up time was 13.0 years. 5-year disease-specific survival (DSS) and progression-free survival (PFS) across the cohort was 72.8% (95% CI 64.8-81.8%) and 68.5% (95% CI 60.2-77.9%).

**Table 1.** Clinical characteristics of the 112 cases characterised by whole exome sequencing

|  | **N / median** | **% / range** |
| --- | --- | --- |
| **Cases** | 112 |  |
| **Age (years)** | 58.5 | 28-88 |
| **BMI** | 25.4 | 18.0-44.0 |
| **Concurrent endometrial cancer<sup>a</sup>** | 19 | 17.0 |
| **Endometriosis<sup>b</sup>** | 39 | 34.8 |
| **Morphology** |  |  |
| G1 EnOC | 69 | 61.6 |
| G2 EnOC | 17 | 15.2 |
| G3 EnOC | 8 | 7.1 |
| WT1 negative tumour of serous or undifferentiated morphology | 18 | 16.1 |
| **Vital status at last follow-up** |  |  |
| Deceased, ovarian cancer | 35 | 31.2 |
| Deceased, other causes | 19 | 17.0 |
| Alive | 58 | 51.8 |
| **Period of Diagnosis** |  |  |
| 1980s | 17 | 15.2 |
| 1990s | 44 | 39.3 |
| 2000s | 32 | 28.6 |
| 2010s | 19 | 17.0 |
| **FIGO stage at diagnosis** |  |  |
| I | 47 | 42.7 |
| II | 39 | 35.5 |
| III | 15 | 13.6 |
| IV | 9 | 8.2 |
| NA | 2 |  |
| **Primary debulking status** |  |  |
| Zero macroscopic RD | 82 | 78.1 |
| Macroscopic RD | 23 | 21.9 |
| NA | 7 |  |
| **Adjuvant chemotherapy stage I/II (n=86)** |  |  |
| Single-agent platinum | 31 | 36.5 |
| Platinum-taxane combination | 17 | 20.0 |
| Other platinum combination | 3 | 3.5 |
| Other chemotherapy regime | 7 | 8.2 |
| No adjuvant chemotherapy | 27 | 31.8 |
| NA | 1 |  |
| **Adjuvant therapy stage III/IV (n=24)** |  |  |
| Single-agent platinum | 14 | 58.3 |
| Platinum-taxane combination | 2 | 8.3 |
| Other platinum combination | 2 | 8.3 |
| No adjuvant therapy | 6 | 25.0 |

BMI, body mass index; G1, grade I; G2, grade II; G3, grade III; EnOC, endometrioid ovarian carcinoma; NA, not available; RD, residual disease
<sup>a</sup>as documented on the Edinburgh Ovarian Cancer Database
<sup>b</sup>as documented on diagnostic pathology report or identified from reviewed archival tissue

**Figure 1:**
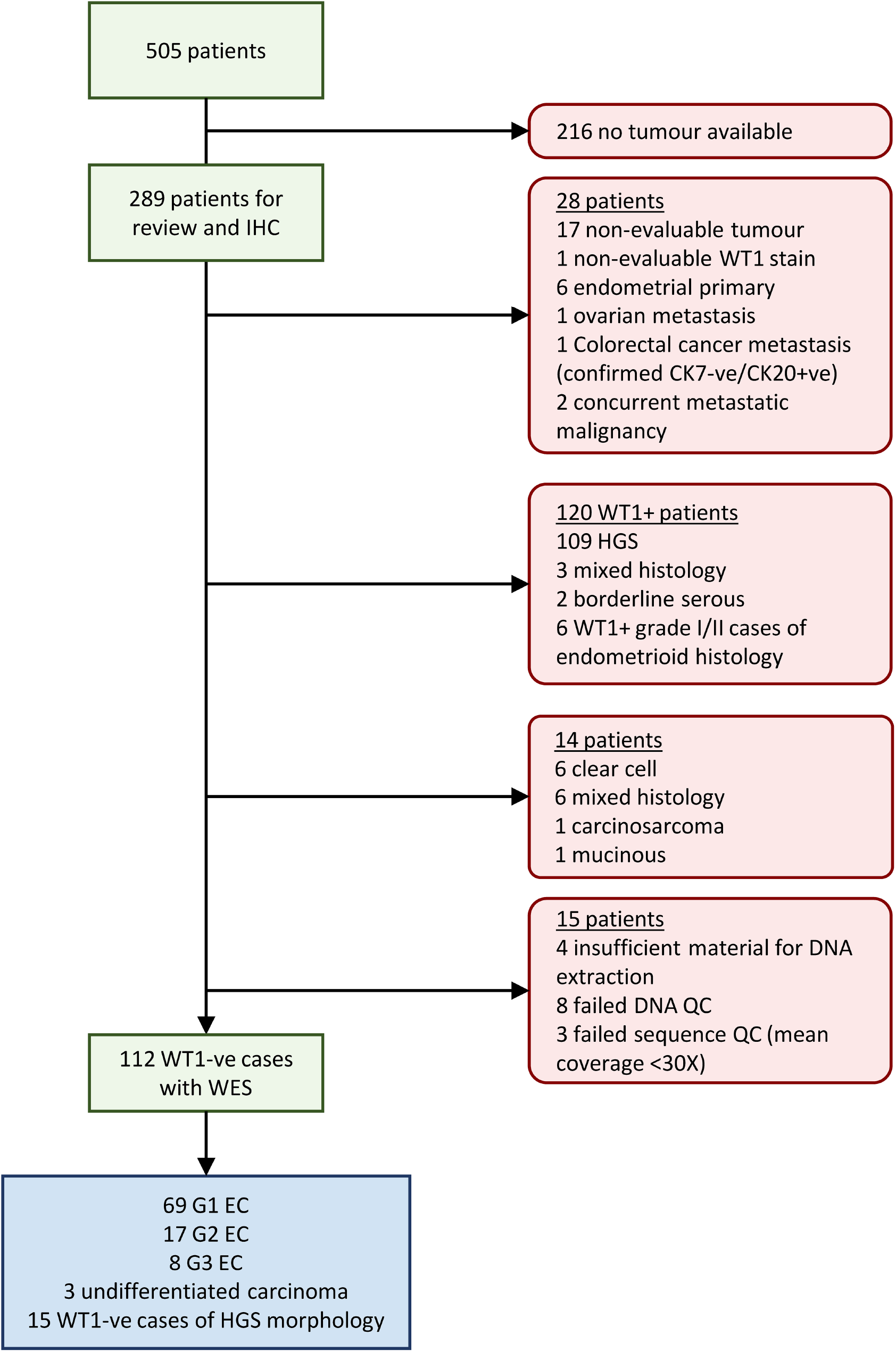
CONSORT diagram demonstrating identification of EnOC cases for whole exome sequencing. IHC, immunohistochemistry; WES, whole exome sequencing; G1, grade I; G2, grade II; G3, grade III; ca, carcinoma

### Molecular landscape of WT1 negative EnOC from whole exome sequencing

The most commonly mutated target genes included *CTNNB1* (48 cases, 42.9%), *PIK3CA* (48 cases, 42.9%), *ARID1A* (40 cases, 35.7%), *PTEN* (33 cases, 29.5%), *KRAS* (29 cases, 25.9%) and *TP53* (29 cases, 25.9%) (figure S1 and S2). Unsupervised hierarchical clustering across the 50 most commonly mutated genes across the cohort revealed mutation of *TP53 and CTNNB1* (*TP53*m and *CTNNB1*m) as the most prominent stratifying events (figure 2). *TP53*m and *CTNNB1*m were largely mutually exclusive, with significant depletion of *CTNNB1*m in the *TP53*m group (P<0.001; co-occurrence in 1 case, 0.9%).

**Figure 2:**
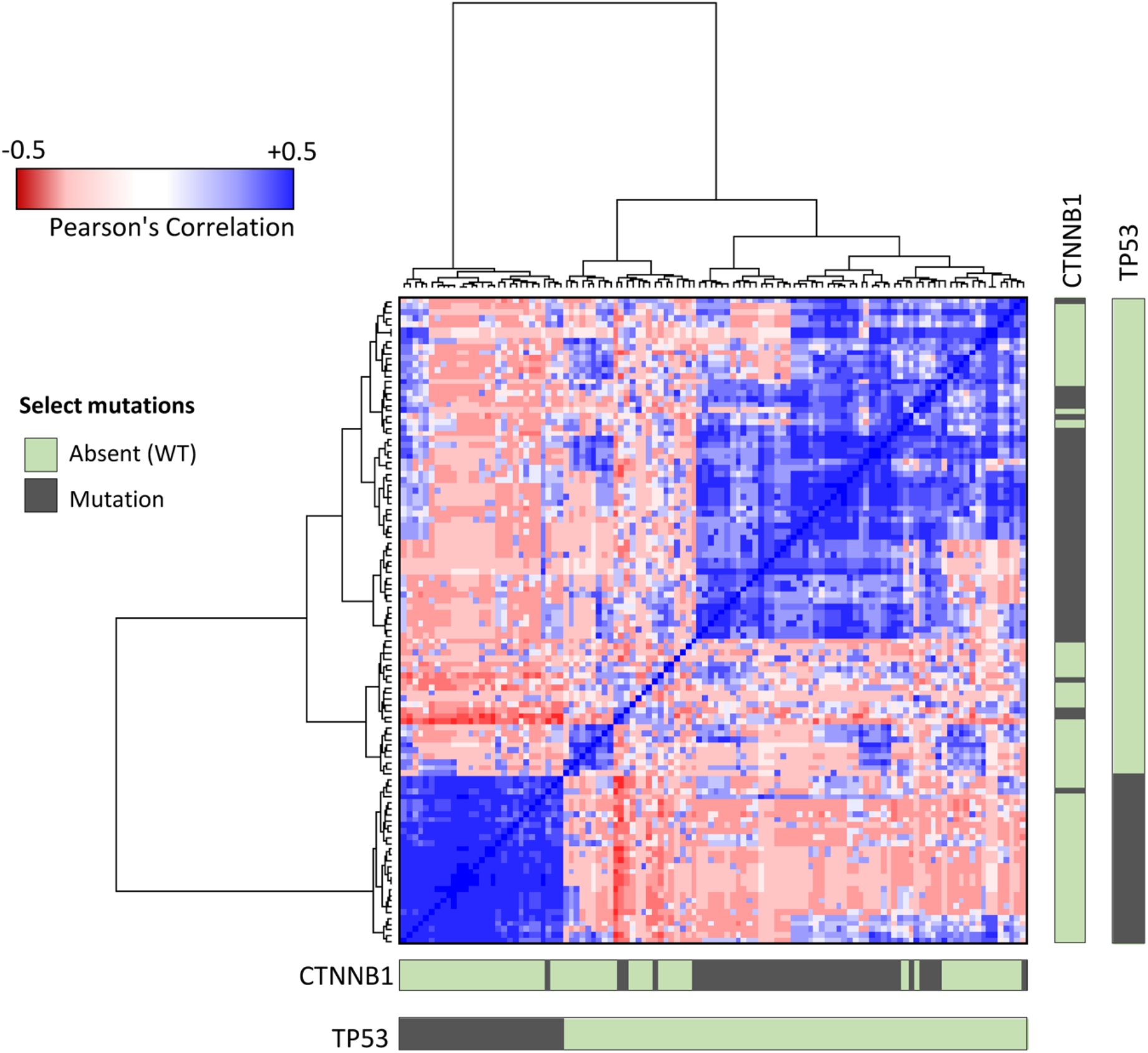
Unsupervised hierarchical clustering of endometrioid ovarian carcinomas using Pearson’s correlation scores based on the mutational status of the most frequently mutated genes. Bars denote major stratifier genes, *CTNNB1* and *TP53*.

Mutation in *SOX8*, a novel target of recurrent mutational disruption in EnOC, was also identified at high frequency (21 cases, 18.8%), alongside other novel targets of mutational events (figure 3A, supplemental figure S3A). There was significant enrichment of *SOX8*m in the *TP53*m group (10/29, 34.5% *SOX8*m in the *TP53*m group vs 11/83, 13.3% *SOX8*m in the *TP53* wild-type (*TP53*wt) group; P=0.025). Events in other genes previously reported as mutated in EnOC or endometrial cancers were identified at lower frequency, including *FBXW7*m (14 cases, 12.5%), *KMT2D/MLL3*m (12 cases, 11%), *APC* (6 cases, 5.4%), *BRCA1/2*m (14 cases, 12.5%), *MTOR*m (7 cases, 6.5%), *PIK3R1*m (10 cases, 8.9%) and *PPP2R1A*m (4 cases, 3.6%) (figure 3A, S1 and S2). Combined analysis of frequently mutated pathways across the total EnOC tumour set identified a large number of mutations across four major oncogenic pathways: PI3K-AKT, WNT, RAS & NOTCH (supplemental figure S4).

**Figure 3:**
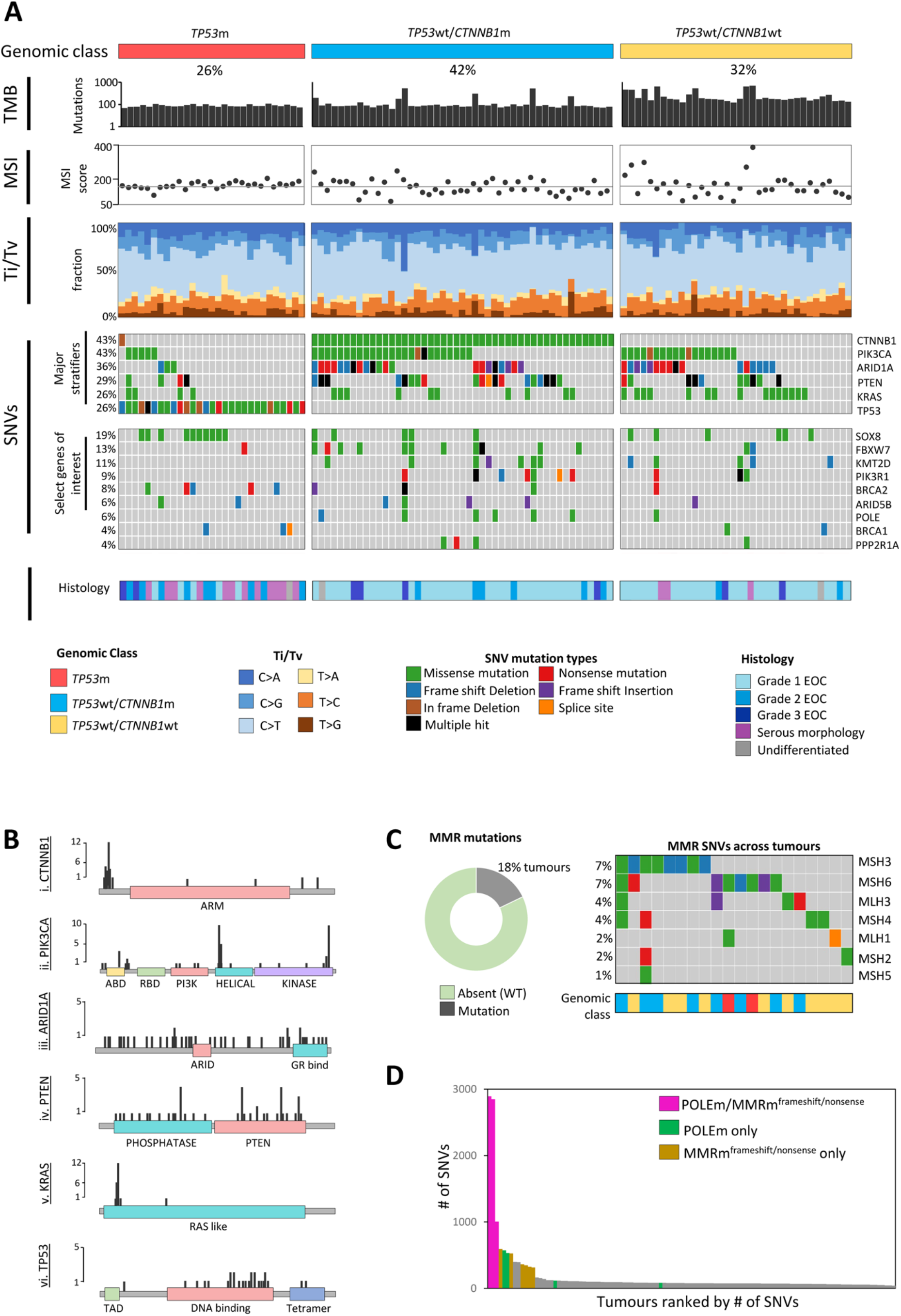
Genomic characterisation of endometrioid ovarian carcinoma. **A**. Whole exome sequencing identifies three major genomic subtypes of endometrioid ovarian carcinoma based on SNV status. TMB is plotted as number of SNVs per sample. Microsatellite instability (MSI) scores are plotted based on INDEL counts and colour coded based on mutations across mismatch repair genes. Molecular signatures for each tumour are plotted as the fraction of transversions and transitions (Ti/Tv). SNVs are displayed as an oncoplot. Grey denotes no mutation. Percentages on the left indicate % of samples with a given mutation. Upper plot shows the six most frequently mutated genes which act as major stratifiers, lower plot shows select genes of interest. **B**. Lollipop plots of five common gene targets of mutation. **C**. MMR mutations across cases. **D**. Relationship between TMB and mutations in *POLE* and/or MMR genes.

### Tumour mutational burden and microsatellite instability in EnOC

A median of 78 variants were detected per sample (range 42-2,894) (supplemental figure S5 & s6A). 10 tumours (8.9%) were considered ‘hypermutated’ (>250 mutations per sample) and 3 (2.7%) were considered ‘ultramutated’ (>1000 mutations) (Figure 3A). Overall analysis of tumour mutational burden (TMB) against TCGA derived datasets places EnOC alongside HGSOC (median mutation count = 72), colonic adenocarcinoma (median = 76) and EnEC (median = 78) (supplemental figure S6B).

Mutations in one or more genes encoding MMR proteins were identified in 20 (17.9%) cases (figure 3C), most commonly in *MSH3, MSH6, MLH3* or *MSH4*. The majority of MMR-mutant (MMRm) tumours were *TP53*wt (18/20 cases, 90.0%). Frameshift or nonsense MMRm were associated with significantly higher microsatellite instability (MSI) scores compared to those containing missense MMRm (median 236 vs 142.5, P<0.001) and MMRwt samples (median 236 vs 162, P<0.001) (figure 3A & supplemental figure S7). There was no significant difference in MSI score between MMRwt cases and those with missense MMRm (P=0.181).

*POLE*m were detected in a minority of tumours (7 cases, 6.3%) with 3 (42.8%) found to occur over a hotspot within the exonuclease domain (Figure 3A & supplemental figure S3B). There was a high frequency of concurrent *POLE*m and MMRm (5 of 7 *POLE*m cases, 71.4%), with significant enrichment for *POLE*m in the MMRm group versus MMR wild-type cases (5/20, 25.0% vs 2/92, 2.2%, P=0.002). Together, the *POLE*m and nonsense/frameshift MMRm cases accounted for majority of high TMB cases (figure 3D). Cases with concurrent *POLE*m and MMRm accounted for all three ultramutated tumours; 8 of the 10 (80.0%) hypermutated tumours contained either *POLE*m or frameshift/nonsense MMRm.

### Mutational spectrum in endometrioid ovarian carcinoma

We observed a bias towards C>T and C>A transversion and transition molecular signatures across the 112 tumours (figure 3A). A shift in signatures was observed in samples harbouring *POLE*m, with a greater proportion of T>G changes, and depletion of C>G and T>A substitutions in this population (supplemental figure S8).

### Tumour complexity and copy number variation

Distribution of per-sample global variant allele frequency (VAF) density and calculation of mutant-allele tumour heterogeneity (MATH) score across the 112 EnOC samples was used to infer tumour complexity (figure 4). *TP53*wt tumours were predominantly low complexity, demonstrating lower MATH scores (median 27.5 vs 54.7, P<0.001) (figure 4B & supplemental figure S9) and fewer discrete VAF peaks (P<0.001) (supplemental table S1) compared to *TP53*m cases.

**Figure 4.**
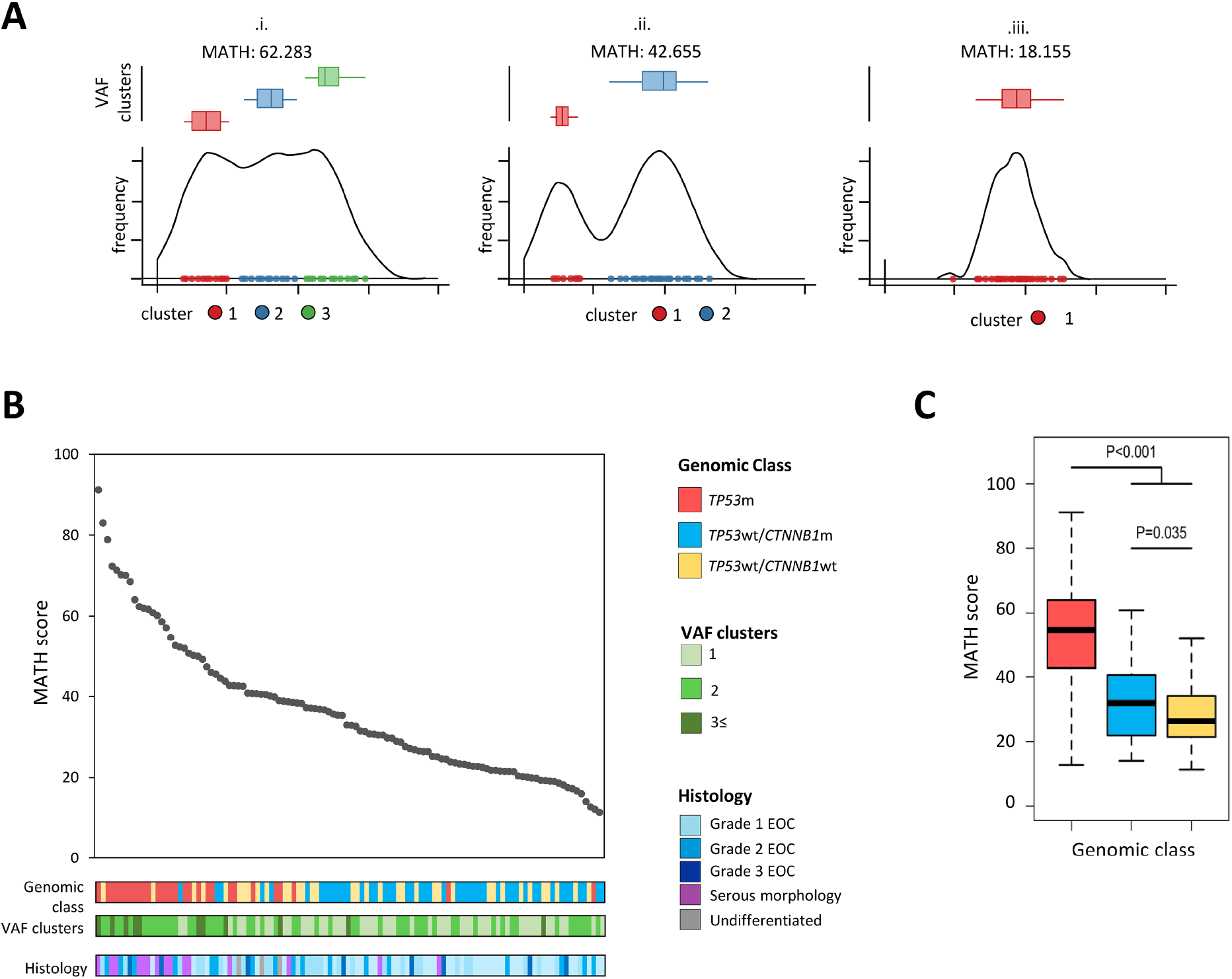
Genomic complexity of endometrioid ovarian carcinomas. **A**. Example density plots for (i) complex, (ii) intermediate and (iii) low complexity tumours. Clusters of VAF scores are shown and summarized within box plots above the density plot. **B**. plot of MATH scores across all 112 tumours, ranked high to low. Bars below show genomic class, number of discrete VAF peaks and histologically defined tumour grade.**C**. Box plot of MATH scores across the three genomic classes.

Analysis of copy number variation (CNV) across the samples revealed differential CNV burden across the molecular subgroups (supplemental figure S10A & s10B), with *TP53*m cases harbouring greater CNV burden compared to *TP53*wt tumours (P<0.0001). The most frequent CNVs across the cohort were CN gain of *ZNF43* (30 cases, 26.8%), *RABA1C* (24 cases, 21.4%) and *AMY1C* (23 cases, 20.5%), and CN loss of *PKNOX1* (42 cases, 37.5%), *CEP68* (29 cases, 25.9%) and *GLB1L* (20 cases, 17.9%) (supplemental figure S10C) with CNV events also occurring over some of the genes identified as containing frequent SNVs (supplemental figure 10D).

### Molecular events in EnOC define clinically distinct subgroups of disease

*TP53*m cases demonstrated significantly inferior survival upon univariable analysis (HR for DSS=4.43, 95% CI 2.27-8.64, Bonferroni-adjusted P<0.001) (figure 5Ai, and table S2), were more likely to be diagnosed at advanced stage (14 of 29 evaluable cases, 48.3% stage III/IV vs 10 of 81, 12.4%; P<0.001) (table S3), less likely to be successfully resected to zero macroscopic residual disease (44.4%, 12 of 27 evaluable cases with macroscopic RD after surgical debulking vs 14.1%, 11 of 78; P=0.003), and demonstrated a trend for greater age at diagnosis which did not meet statistical significance (median 61 vs 57 years, P=0.063). Multivariable analysis accounting for patient age, stage at diagnosis and extent of residual disease following primary cytoreduction identified *TP53* status as independently associated with survival (P=0.031 for DSS; P=0.011 for PFS) (table S4 and S5, figure S11). The *TP53*m group demonstrated significant depletion of cases with concurrent endometrial cancer diagnosis (3.4%, 1 of 29 *TP53*m vs 21.7%, 18 of 83 *TP53*wt, P=0.022).

**Figure 5.**
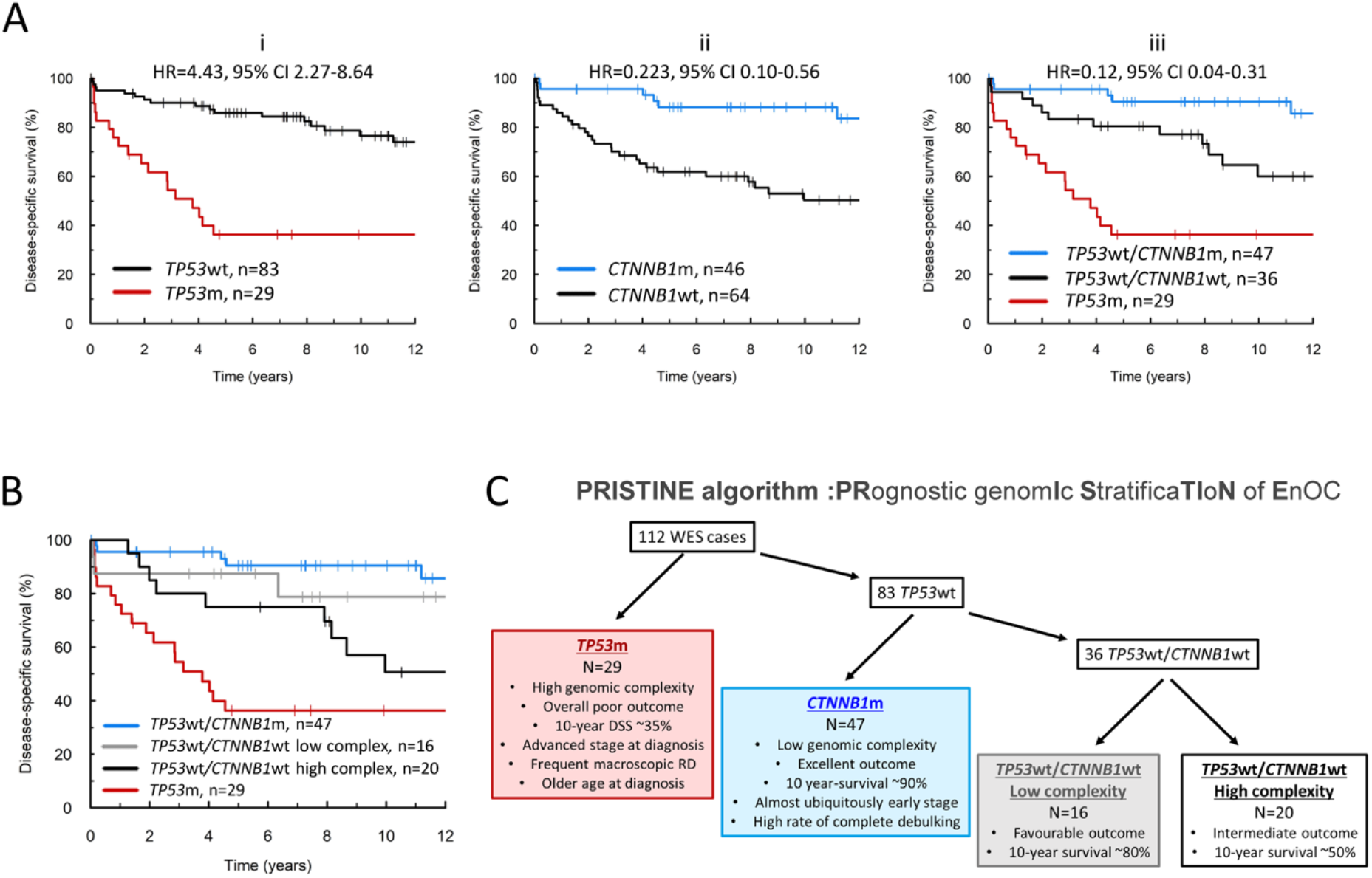
Disease-specific survival of molecularly-defined endometrioid ovarian carcinoma (EnOC) subtypes. (A) Disease-specific survival based on *TP53*m (i), *CTNNB1*m (ii) or combined mutational status (iii). (B) Disease-specific survival based on the PRISTINE algorithm for EnOC classification. (C) Summary of the PRISTINE algorithm for molecular subtyping in EnOC.

By contrast, cases with *CTNNB1*m were almost ubiquitously stage I/II at diagnosis (89.1%, 41 of 46 evaluable cases) and debulked to zero macroscopic RD (87.0%, 42 of 46 evaluable cases), with markedly favourable outcome (HR for DSS=0.23, 95% CI 0.10-0.56, Bonferroni-adjusted P=0.010) (table S2) which was significant upon multivariable analysis (P=0.017 for DSS; P=0.006 for PFS) (figure 5Aii, supplemental s6 and s7). *CTNNB1*m was significantly associated with favourable outcome specifically in the context of *TP53*wt cases (HR for DSS 0.30, 95% CI 0.11-0.87), and *TP53* wild-type/*CTNNB1*m cases were less genomically complex versus their *TP53*wt/*CTNNB1*wt counterparts (median MATH score 36.3 vs 31.9, P=0.035) (figure 4C).

Greater tumour complexity was associated with inferior survival when defined by the number of VAF peaks (P=0.020 for DSS) or continuous MATH score (P<0.001 for DSS) across the cohort (supplemental table S7). Moreover, MATH score at a threshold of the median score in low complexity tumours (with a single VAF peak, figure 4Aiii), provided additional meaningful prognostic information specifically in the absence of *TP53*m or *CTNNB1*m (figure 5B, figure S11). Together, these data support a clinically meaningful classification system driven firstly by *TP53*m status, then *CTNNB1*m status, with genomic complexity providing further discrimination of outcome within the *TP53*wt/*CTNNB1*wt population; we define this step-wise molecular classification system as the **PRISTINE algorithm :<u>PR</u>**ognostIc genom<u>**I**</u>c **<u>S</u>**tratifica**<u>TI</u>**o**<u>N</u>** of **<u>E</u>**nOC (figure 5C).

## Discussion

The molecular landscape of EnOC is poorly defined, particularly in high grade cases, due to under-investigation and historic misclassification of HGSOC as high grade EnOC in older studies. WT1 negativity has emerged as an important discriminator of high grade EnOC from HGSOC, which displays morphological similarities^4, 23, 24, 25, 26^. To our knowledge, this is the largest report of genomically characterised EnOC to date, utilising routine WT1 IHC to exclude pseudo-endometrioid HGSOCs carcinomas that have contaminated previous studies of this OC type.

In line with previous sequencing studies in small cohorts of EnOC, we identified a high mutation rate of genes known to be perturbed in EnEC, the most frequent of which were *CTNNB1, PIK3CA, PTEN, ARID1A, KRAS* and *TP53* ^7, 10, 21, 27, 28^. Our EnOC cohort demonstrated a similar rate of *TP53*m to the TCGA study of EnEC^10^; by contrast, the mutation rate of *PTEN* was markedly lower.

We demonstrate that EnOC tumours contain a moderate tumour mutational burden with respect to other cancer types, with a small proportion hyper-or ultra-mutated in nature (11.6%, 13 of 112). MMR deficiency due to mutations or methylation in the MMR genes results in microsatellite instability, and loss of MMR protein expression has previously been demonstrated in approximately 7-14% of EnOC^29, 30, 31, 32, 33^. As in EnEC, we identify a subgroup of EnOC harbouring MMR mutations, with a similar overall MMR mutation rate as reported previously in EnOC^29, 30, 31, 32, 33^. We demonstrate that cases with truncating MMRm (nonsense or frameshifting) demonstrate MSI and that these cases account for many of the samples demonstrating high TMB. Moreover, mutation of *POLE* was rare in this cohort (6.3% of cases), there was significant enrichment for this event in the MMRm population, and concurrent mutation of *POLE* and MMR genes was present in all three ultramutated tumours. Moreover, *POLE*m cases demonstrated a distinct mutational profile. However, unlike in EnEC^10^, *POLE*m was not associated with a significant survival benefit, though the power of these analyses was severely limited. This low *POLE*m rate is consistent with previous reports in EnOC^34, 35^.

Collectively, the data presented here identify *TP53*m EnOC as a distinct clinical and biological subtype of EnOC. *TP53*m cases demonstrated higher levels of copy number aberrations, greater tumour genomic complexity, higher rate of advanced stage at diagnosis, inferior rate of complete macroscopic tumour resection, and overall poor clinical outcome. This is consistent with the poor outcome reported in EnEC harbouring *TP53*m^10^, is reminiscent of HGSOC^11^, and is in line with several studies of EnOC^27, 36^. In particular, the study performed by Parra-Herran et al^27^ applied the PROMISE algorithm, a surrogate of the EnEC molecular classifier, to a cohort of WT1 negative EnOC and found the group with aberrant p53 expression to have the worst survival. In our cohort, *TP53*m cases also represented those least likely to demonstrate concurrent endometrial cancer.

While the copy number and survival profile of our *TP53*m EnOC group provides one rationale for reclassification of these tumours as HGSOC, the high frequency (48.3%, 14 of 29) of MMRm or classic EnOC mutations (*CTNNB1, PTEN, ARID1A, KRAS* or *PIK3CA*), lack of WT1 expression (in all cases) and high rate of early stage diagnosis in this cohort (51.7%, 15 stage I/II) form a compelling argument that these represent true EnOC. Only 9 cases (8.0% of our cohort) represented feasible candidates as possible true WT1-negative HGSOCs (MMR, *PTEN, CTNNB1, PIK3CA, ARID1A, KRAS* wild-type tumours with *TP53*m and diagnosed at stage III/IV), which are recognised as a rare phenomenon (≤5% HGSOC cases)^23^.

Conversely, *CTNNB1*m – which appears mutually exclusive with *TP53*m – is associated with low genomic complexity, early-stage disease that is easily debulked to zero macroscopic RD and demonstrates excellent clinical outcome. This is in contrast to findings in low grade early stage EnEC associating *CTNNB1*m with a greater chance of recurrence^37^. Within our EnOC cohort, *CTNNB1*m status was also associated with superior outcome specifically in the context of *TP53*wt tumours, suggesting clinical impact independent of its anti-correlation with *TP53*m. These data support the notion of a tiered classification by *TP53*m and *CTNNB1*m status. The outcome of the remaining *TP53*wt/*CTNNB1*wt patients, who demonstrate an overall intermediate survival profile and moderate genomic complexity, can be further resolved by tumour complexity score. *TP53* status, *CTNNB1* status and tumour complexity can therefore be used in succession within the PRISTINE algorithm for defining molecular subtypes with markedly differential clinical outcome and clinicopathological features.

The high rate of genomic disruption in *CTNNB1, KRAS, PTEN* and *PIK3CA* suggests that inhibitors of the WNT, MAPK and PI3K pathways represent agents with potential clinical utility in EnOC treatment. Efforts to identify novel therapeutic strategies should focus on cases with greatest unmet clinical need, namely *CTNNB1*wt cases. In particular, *TP53*m cases represent those where further treatment options are urgently required to improve outcome, and we identified potentially clinically actionable mutations in a large proportion of these cases (13.8% with *KRAS*m, 27.6% with *PTEN*m/*PIK3CA*m, 27.6% with *BRCA1/2*m).

Finally, we identify *SOX8* as a potential novel target of mutational disruption in EnOC. SRY-related high mobility group (HMG) box (*SOX)* genes encode a family of transcription factors which act as critical regulators of cellular programming and are frequently altered in many cancers^38^. Interestingly, analysis of TCGA data reveals that *SOX8*m occur at low frequencies in uterine cancers (2.64% in uterine corpus endometrial carcinoma, 1% in uterine carcinosarcoma) as well as in colonic adenocarcinoma (2.25%) (data from the TCGA data portal^39^). As recent studies suggest that *SOX8* in part regulates the activity of genes associated with the WNT/β-catenin pathway, a commonly mutated pathway in EnOC, mutation of this gene may impact on classically defined EnOC pathways through this route^40^. Genomic disruption of *SOX8* therefore represents a novel candidate mechanism by which EnOCs frequently activate the WNT/β-catenin pathway; given the frequent co-occurrence of *SOX8*m and *TP53*m in our cohort (34.5% SOX8m in the TP53m group), selection bias against true high grade EnOCs in previous studies -leading to depletion of *TP53*m cases in those cohorts -may well explain why *SOX8*m has not previously been identified as a common genomic event in EnOC.

In summary, we have demonstrated that EnOC is a molecularly heterogeneous disease, comprising molecular subtypes of disease which we define here using the PRISTINE algorithm. These subtypes demonstrate differential clinical outcome and clinicopathological features. In particular, our study highlights *CTNNB1*m and *TP53*m as markers of biologically distinct subtypes of EnOC with contrasting clinical behaviour. These markers have the potential to inform future prognostication and molecular stratification within EnOC. Gene sequencing of *TP53* and *CTNNB1*, or IHC directed at their respective gene products, represent mechanisms by which these findings could readily be translated into routine risk-stratification of newly diagnosed cases. EnOCs demonstrating absence of *CTNNB1*m and/or presence of *TP53*m represent patients with greatest unmet clinical need, many of whom harbour activating mutations in pathways that represent opportunities for therapeutic intervention with molecular agents. Investigating the clinical efficacy of inhibitors targeting the MAPK/RAS, WNT and PI3K pathways in this context has the potential to identify agents that will improve EnOC patient survival.

## Methods

### Ethical approval

Ethical approval for the use of human tissue specimens for research was obtained from South East Scotland Scottish Academic Health Sciences Collaboration (SAHSC) BioResource (reference: 15/ES/0094-SR494). Correlation of molecular data to clinical outcome and clinicopathological variables in ovarian cancer was approved by NHS Lothian Research and Development (reference 2007/W/ON/29).

### Pathology review

505 patients diagnosed with OC between August 1968 and May 2014, and whose pathology reports contained the term ‘endometrioid’, were identified through the Edinburgh Ovarian Cancer Database (figure 1); tumour material was available for 289 cases^41^. Chemotherapy naïve tumour from the primary site was selected where available. Pathology review was conducted as per WHO 2014 classification, including immunohistochemistry (IHC) for WT1 in every case (Supplementary Methods Section 1A), by an expert gynaecological pathologist (CSH). The presence of endometriosis was recorded from the reviewed slides or pathology report.

Cases with non-interpretable morphology, non-evaluable tumour and cases representing metastases from primary endometrial cancer, as defined by WHO criteria, were excluded. Ovarian metastases, WT1 positive tumours, carcinosarcomas and carcinomas of clear cell, mucinous or mixed histology were also excluded (figure 1). IHC for CK7 (Leica, Clone RN7, 1:100 dilution) and CK20 (Leica, clone KS20.8, 1:50 dilution) was performed to exclude colorectal adenocarcinoma metastases (Supplementary Materials Section 1B).

### Clinical data

Baseline characteristics and outcome data were extracted from the Edinburgh Ovarian Cancer Database, wherein the diagnostic, treatment and follow-up data for every ovarian cancer patient treated at the Edinburgh Cancer Centre is prospectively entered as part of routine care. Disease-specific survival (DSS) was calculated from the date of pathologically confirmed OC diagnosis. Progression-free survival (PFS) was recorded as the duration between the date of diagnosis to the date of first radiological progression or recurrence, or death from EnOC.

### DNA extraction

H&E-stained slides were marked by an expert gynaecological pathologist (CSH) to identify tumour areas suitable for macrodissection in order to enrich for tumour cellularity. DNA extraction was performed using the QIAamp DNA FFPE Tissue Kit (Qiagen) and Deparaffinization Solution according to the manufacturer’s instructions.

### Whole exome sequencing

Exome capture was performed using the Illumina TruSeq Exome Library Prep kit (supplementary materials section 2A) and WES was performed on the Illumina NextSeq 550 (Illumina, Inc., San Diego, CA, USA). The median per-sample on-target coverage in the successfully sequenced samples was 89.5X (range 36X-289X). Data were aligned to the GRCh38 human reference genome using bwa-0.7.17^42^, duplicates marked and base quality scores recalibrated with the GenomeAnalysisToolkit (GATK) v4^43^ in the bcbio 1.0.6 pipeline (Supplementary Materials Section 2B).

### Variant calling and classification

Variant calling was performed using a majority vote system from three variant caller algorithms: VarDict^44^, Mutect2^45^ and Freebayes^46^. Filtering for C>T (FFPE artifacts) and G>T (oxidation artifacts) was applied using GATK (CollectSequencingArtifactMetrics and FilterByOrientationBias). Variants associated with low sequence depth (<20X) or low variant allele frequency (<10%) were removed.

Common variants were excluded using the 1000 genomes and ExAC reference datasets; known pathogenic and benign variants were flagged using ClinVar^47^, and remaining variants were filtered to remove likely non-functional variation using the Polymorphism Phenotyping (PolyPhen)^48^ and Sorting Intolerant from Tolerant (SIFT)^49^ functional prediction tools (Supplementary Materials Section 2C).

Microsatellite instability (MSI) score was assessed as the number of short insertions or deletions (InDels) detected in a given sample. TMB was defined as the number of mutations present in a given tumour following the aforementioned filtering steps. TMB across other cancer datasets in The Cancer Genome Atlas (TCGA) were contrasted against those in our EnOC datasets^50^.Transitions and transversions were calculated using the titv function in maftools^51^.

Unsupervised analysis was performed across the top 50 differentially mutated genes within the total tumour dataset represented as a binary matrix (0, wild-type; 1, mutant). Samples were clustered by Euclidian distance and Ward’s linkage based on the overall Pearson correlation score of these binary signatures. Heat maps were drawn in R using the ggplot package. Supervised mutational analysis was performed using the most commonly mutated genes across sequenced samples. Genomic events in these genes and overall tumour mutational burden analysis were visualised using the R package maftools^51^. Pathway analysis was carried out using the OncogenicPathways function^52^.

### Tumour complexity scoring

Tumour genomic complexity was assessed by VAF density using the inferHeterogeneity function in the R package maftools^51, 53^ (supplementary materials section 3). Resulting MATH scores represent the width of the VAF distribution; specimens of low complexity with a single driver event and associated outgrowth demonstrate fewer VAF peaks with a lower MATH score. Conversely, highly complex tumours with multiple driver events, branched evolution and multiple subclonal populations demonstrate multiple VAF peaks and higher MATH score. With the PRISTINE algorithm, the *TP53*wt/*CTNNB1*wt group were split into higher and lower complexity using MATH score, at a cutoff equal to the median MATH score of low complexity tumours (with a single VAF peak, figure 4Aiii).

### Copy number variation (CNV) detection

Copy number analysis was performed using GeneCN pipelines using Bio-DB-HTS version 2.10 (see Supplementary Materials Section 4) to identify regions of significant copy number gain or loss (CN score >5 standard deviations from reference, P<0.05) using the pooled *TP53*wt samples as a reference population.

### Statistical analysis

Statistical analyses were performed using R version 3.5.1. Comparisons of continuous data were made with the Mann Whitney-U test or T-test, as appropriate. Median follow-up time was calculated using the reverse Kaplan-Meier method. Survival analysis was performed using the Cox proportional hazards regression analysis in the Survival package^54^. Multivariable analyses accounted for FIGO stage, patient age at diagnosis, decade of patient diagnosis and extent of residual disease following surgical cytoreduction. Comparisons of frequency were performed using the Chi-squared test or Fisher’s exact test, as appropriate. Correction for multiplicity of testing was performed using the Bonferroni method where appropriate.

## Data Availability

The data used to generate the analyses presented here are currently being submitted to the European Genome-phenome Archive (EGA) and once data upload completed will be made available upon request to our data access committee. For more information please see https://ega-archive.org/access/data-access

## Acknowledgements

We extend our thanks to the patients who contributed to this study and to the Edinburgh Ovarian Cancer Database from which the clinical data reported here were retrieved. We thank the Edinburgh Clinical Research Facility, Western General Hospital, Edinburgh, UK for their support with the high throughput sequencing described here. We are grateful to the NRS Lothian Human Annotated Bioresource, NHS Lothian Department of Pathology and Edinburgh Experimental Cancer Medicine Centre for their support.

## Data availability

The data used to generate the analyses presented here are currently being submitted to the European Genome-phenome Archive (EGA) and, once data upload is completed, will be made available upon request to our data access committee. For more information please see https://ega-archive.org/access/data-access

## Author contributions statement

RLH contributed conceptualisation, data analysis, visualisation, manuscript writing; JPT contributed data analysis, visualisation, manuscript writing; BS contributed conceptualisation, cohort collation, data collection, manuscript writing; MC contributed project management, tissue access, manuscript review and editing; AMM contributed data analysis, manuscript review and editing; TR, CB and YI contributed data collection, manuscript review and editing; IC contributed tissue processing, manuscript review and editing; MM and FN contributed access to patients/tissue, manuscript review and editing; AO contributed supervision, manuscript review and editing; CAS contributed supervision, manuscript review and editing; CG contributed: conceptualisation, access to patients/tissue, supervision, funding acquisition, manuscript review and editing; CSH contributed: conceptualisation, data collection, supervision, manuscript review and editing.

